# Quality of life associated with clinical and sociodemographic determinants in the postpartum period of Mexican women living with HIV. A cross-sectional study

**DOI:** 10.1101/2025.08.07.25333257

**Authors:** Mauricio Domínguez-Castro, Alicia Ramírez-Ramírez, Noemí G. Plazola-Camacho, Miroslava Avila-García, Margarita López-Martínez, Norah Lucky Katende-Kyenda, Ismael Mancilla-Herrera, Diana M. Soriano-Becerril, José Romo-Yáñez, Carmen S. García-Romero, Ricardo Figueroa-Damián, Jessica Hernández-Pineda

## Abstract

The HIV epidemic remains a major public health challenge, particularly for women living with HIV (WLWH) who face vulnerabilities during pregnancy and motherhood. Addressing factors influencing quality of life (QoL) in this population is vital for improving health outcomes and guiding public health strategies. A cross-sectional study of 75 postpartum WLWH was conducted between 2020-22, using the WHOQoL-HIV-BREF instrument and statistical analyses to evaluate QoL and its associations with socio-demographic and clinical factors. Postpartum WLWH experienced overall medium QoL, with the psychological domain scoring lowest and physical health and independence scoring highest. Nearly half reported poor QoL, influenced by socio-demographic factors and CD4 counts. The WHOQoL-HIV-BREF instrument demonstrated strong reliability, highlighting the multifaceted challenges these women face in maintaining QoL during postpartum. HIV/AIDS affects QoL in postpartum WLWH, with antiretroviral therapy adherence and optimal CD4 levels improving outcomes. These findings support including QoL as a marker of success in WHO strategies.

## Introduction

The HIV epidemic continues to be one of the most significant public health challenges after more than 40 years of its emergence. To date, people living with HIV (PLWH) face challenges related to HIV infection, even when they have had sustained antiretroviral therapy (ART) and they achieve suppression of viral load (VL) of HIV [1–3]; two of the three 95-95-95 important UNIADS targets for eradicating HIV pandemic [4–6]. For instance, from the clinical point of view, PLWH faces chronicity of HIV infection and the effects of long ART treatment and their adherence, as well as the non-infectious comorbidities associated with both [7]. Therefore, from the social point of view, PLWH faces persistent stigma and discrimination [2]. Hence nowadays, clinical and social factors importantly affect their Quality of Life (QoL) [8,9]. Particularly, in specific highly vulnerable groups, as a woman living with HIV (WLWH) [10] in some stage of their life such as pregnancy, postpartum and motherhood, exist additional factors that can further affect their QoL, such as, pain and physical discomfort [11] while the environment, social relationships, and personal beliefs may lead to inherent vulnerability, stigma, discrimination, and an awful perception of their health status, and as consequently their QoL decreases, with negative consequences, such as poor care or less attention [12].

It is known that even in non-complicated perinatal period important physical and emotional changes occurs, and factors such as age, parity, social and economic issues, obstetric complications, obesity, history of alcohol dependence, sleep difficulties, stress, anxiety and postnatal depression affect the mother QoL [13,14]. Furthermore, in studies that assess QoL in mothers with chronic disease, their child also had a worse QoL [15,16]. Another study reports to assess health-related HR-QOL in mothers-to-child pairs with low-income, in which their children had worse and behaviour disorder than children from the general population [17]. There were significant associations of maternal postpartum health, both mental and physical, with the health of their child [16]; therefore, the QoL of mother could determine the QoL of her children as they grow up. In this regard, we emphasise that the traditional approach to health has been based primarily on the clinical point of view, based on the detection, treatment and cure of disease. However, health is not simply the absence of disease; it affects the entire life of each patient, making this approach insufficient. Assessment of QoL could be an indicator for self-perception of well-being and health status, it also includes the terms of QoL. QoL had several definitions and instruments to assess it [18]. Furthermore, according to experts, the assessment of QoL should be evaluated with specialised and validated instruments for a population, especially if it involves a population with chronic diseases [19,20]. According to the World Health Organisation (WHO), QoL is the perception that a person has of their position in life, in the context of the culture and value system in which they live, and concerning their objectives, expectations, standards and concerns, and provides a QoL instrument for the PLWH [8].

In this order of ideas, focusing in WLWH represent 53% of all PLWH [21], and most of them are of reproductive age, implying an increased risk of becoming pregnant and exposing HIV to their newborns [22] as well as, considering that each year there were 1.2 million (950,000–1.4 million) pregnant WLWH, and of those, 84% (72–98%) received antiretroviral therapy (ART) to prevent mother-to-child transmission (MTCT) [22]. Expert groups on HIV/AIDS had proposed including a fourth -95 target (%), focussing on the evaluation of the QoL in PLWH as a vision of therapeutic success [8] and considering it as a key component of public health [23] and providing the management strategies, leading to the establishment of new health policies in PLWH, improving the medical care of vulnerable groups such as WLWH. Therefore, aligning with important United Nations Sustainable Development Goals, particularly within the context of countries facing significant challenges related to social inequalities, are necessary. In Latin America (LATAM), QoL among pregnant or postpartum WLWH have made significant progress, but there are still some important gaps that could be addressed in future research, such as QoL beyond the medical perspective, more exploration is required of aspects such as emotional impact, social stigma and support networks and which characteristics of their population influences these factors to understanding how women navigate their condition and motherhood. The objectives of this study were to describe the QoL during postpartum of Mexican WLWH and to determine its association with sociodemographic and physiopathological factors.

## Materials and methods

### Study design

An observational, cross-sectional study was conducted at the Instituto Nacional de Perinatología (INPer) in Mexico City, a renowned Institution for pregnancy care in WLWH. The study was reviewed and registered by the Ethics and Research Committees of the Institute with number 212250-3120771 and was carried out between December 09^th^, 2020 and June 17^th^, 2022. The sample size for this stage was calculated to include 28 women using a probability finite sample for qualitative variables (Z^2^ = 1.96, p=0.5, d^2^= 5%), after delivery, women were referred to their urban medical center. It just came back only for their baby’s follow-up. We considered 50% losses in recruitment during the intervention in postpartum, so the final sample size calculated was 42.

### Women enrolment

Pregnant WLWH receiving ART treatment, over 18 years old, with prenatal care at the INPer were invited to participate. Those who did not agree to participate received their medical care as usual. The participants who accepted signed a written informed consent; their anonymity and privacy were protected. After delivery, participants returned to our facilities in the postpartum period for a clinical history that included sociodemographic, nonpathological diseases, other infectious diseases, obstetric history, sexual behaviours and QoL instrument explained below.

### Quality of life assessment

QoL was assessed using the Spanish version of WHOQoL-HIV-BREF [24] a short version of the WHOQoL-HIV instrument designed for people living with HIV (PLWH), which has been validated in Spain and Latin America (LATAM) [15–17]. The HIV-specific items extracted from WHOQOL-HIV long form were translated and integrated into the Spanish version of WHOQoL-BREF to complete the 31 items of the WHOQoL-HIV-BREF instrument [24], ensuring the instrument’s relevance and reliability in our study.

This instrument measures the QoL through a 31-item scale grouped into six domains as follows: 4-items on Physical health, 5-item on Psychological health, 4-items on level of independence, 4-items on Social relationships, 8-items on Environment, 4-items on Spirituality/personal beliefs and 2-items measure the total or general QoL through the rate of perception of QoL and satisfaction with their health [18]. Individual items are rated on a Likert scale where 1 indicates low/negative perceptions, and 5 indicates high/positive perceptions. Items that ask about negative perceptions and experiences are reverse-coded for scoring. Therefore, higher scores for all items indicated better QoL. The average score for each domain was multiplied by four, producing domain scores ranging from 4 to 20, where 4 means the lowest QoL and 20 is the highest [24]. Considering the mean total QoL score obtained as a cut-off point, the participants’ perceptions of QoL were classified as poor, medium, and high QoL for this study.

The modifications to the instrument consisted of 11 questions in addition to their standard sociodemographic information: type of employment, monthly income, and residence in a rural or urban zone. Additionally, information about clinical data on comorbidities, CD4 count, viral load, type of ART, dosage, and adherence. Eight specific questions related to gestational age, postpartum time, perception of complications in these periods, and support from social networks were added. Our instrument was validated by five HIV specialist partners, four Spanish speakers, and one English-Spanish speaker (**S1 File**). Patients identified as having physical or psychological problems during the intervention were referred to the specialty for additional medical care.

### Statistical Analysis

Means and standard deviations for continuous variables were reported. For categorical variables, the data are presented as frequencies and percentages. The Jarque-Bera test was performed to determine the normality of the data obtained, and then nonparametric analysis was applied. Spearman’s correlation coefficient was used to determine the association of QoL instrument scores with demographic, nonpathological history, infectious history and ART management, obstetric history, and sexual behavior variables. U-Mann Whitney and Kruskal Wallis tests were performed; Dunn’s and Bonferroni’s post-hoc analysis tests were used. A statistical significance of *P*≤ 0.05 was considered in all cases. The Cronbach index (α) was calculated to determine the internal consistency or reliability of the WHOQoL-HIV-BREF instrument. From the α-index range (0-1), the acceptable benchmark value considered was 0.7, which is acceptable from 0.7 to 0.79. Statistical analyses were performed using GraphPad Prism v7 (Dotmatics, UK).

## Results

### Characteristics of the study population

The sample consisted of 75 postpartum-WLWH undergoing different ART, with an average age of 29±5.4 years. Upon admission to INPer, most were in the third trimester of pregnancy, averaging 32±8 gestational weeks. Among these women, 76% had completed nine years of education, 47% cohabited with their partners, 69% were housewives, and 92% resided in urban areas, as presented in **Table 1**. Monthly incomes ranged from USD $29.2 to USD $1755.00, placing 97% in the low-income classification according to the National Institute of Statistics and Geography (INEGI, Mexico) [25].

**Table 1.**
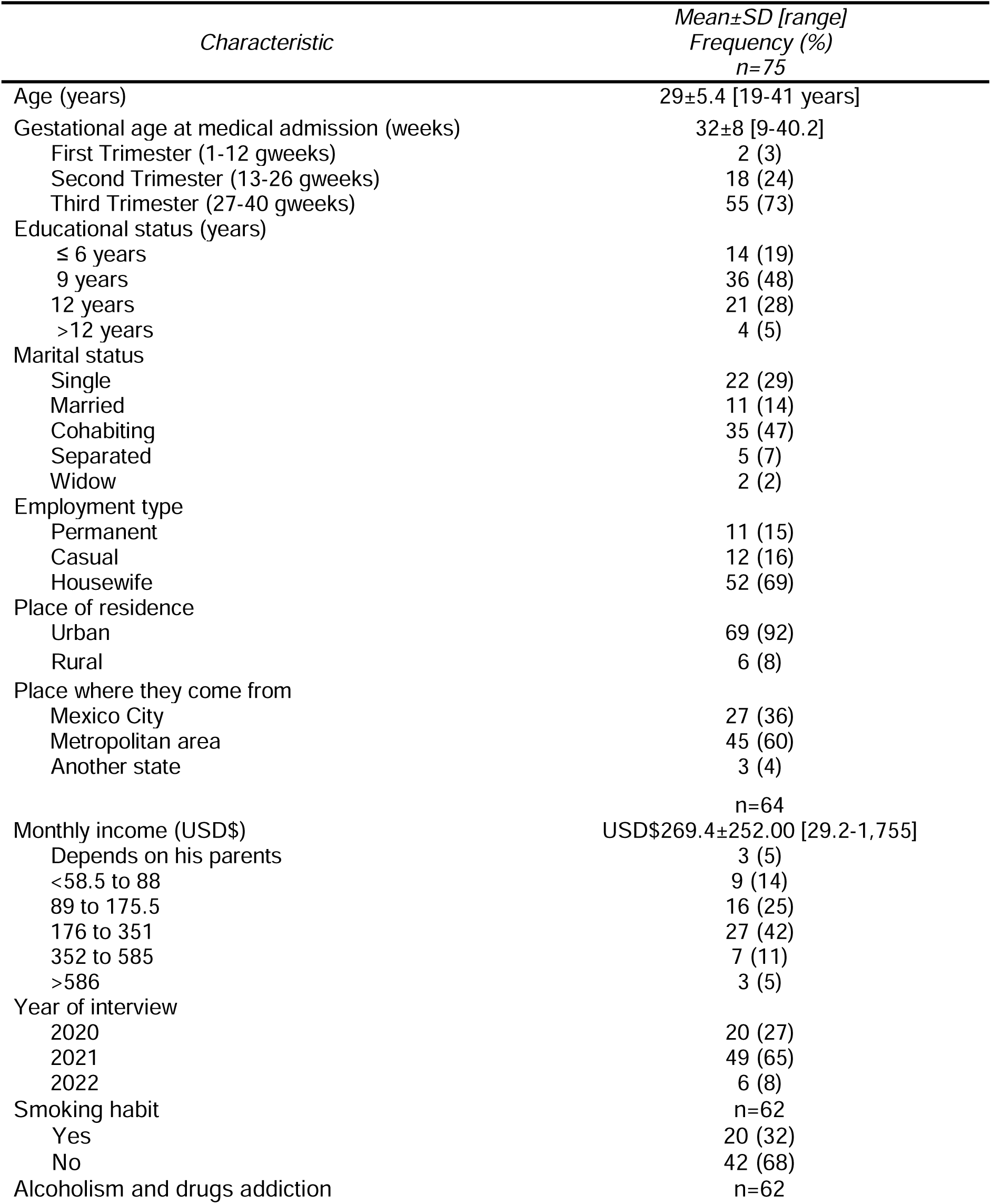
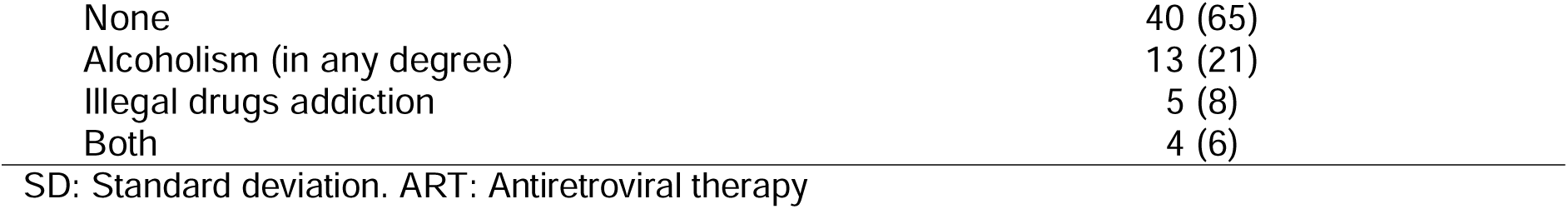
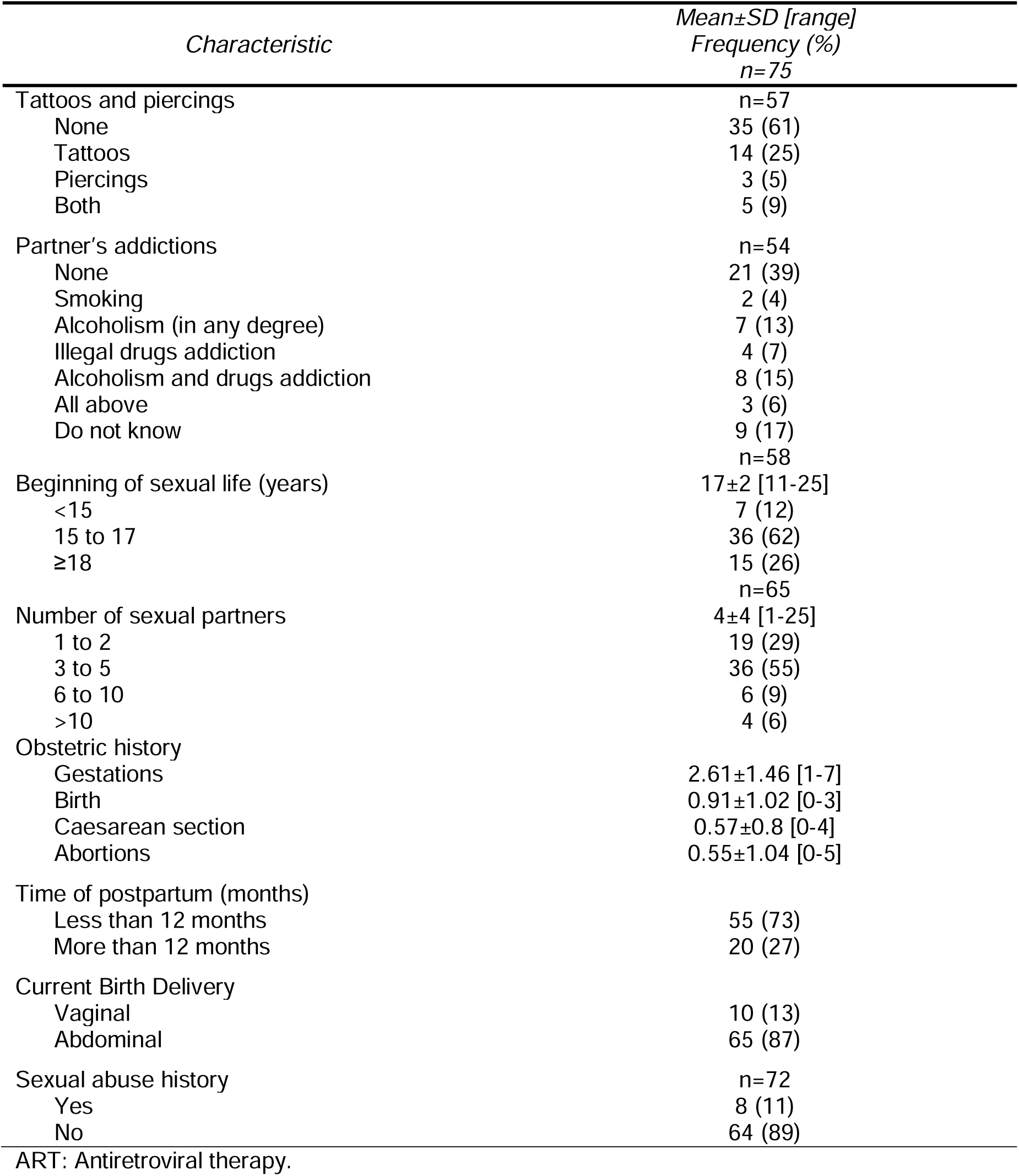
Demographic characteristics, obstetric history and sexual behavior of women living with HIV in postpartum using ART

Regarding drug use during their current pregnancies, participants denied current consumption of alcohol, tobacco, or illegal substances. However, they reported alcohol consumption in 21%, illegal drug use in 8%, and minimal cases of combined use (6%) histories, as detailed in **Table 1**.

Concerning the partner’s drug addiction, participants denied knowledge of their current partner’s substance use (17%), while reporting smoking (4%), alcoholism (13%), drug addiction (7%), combined alcohol and drug use (15%), and a mix of alcohol, smoking, and drugs (6%) among those aware of it. Concerning tattoos and piercings, 25% had tattoos, 5% had piercings, and 9% reported both (**Table 1**).

In terms of sexual behavior, most of the participants initiated sexual activity in late adolescence, with a history of 3 to 5 sexual partners until the study period. Unfortunately, 11% reported having been sexually abused. They were multigravida, delivered by Caesarean section, and reported pregnancy loss (0.55±1.04) based on their gynaecological and obstetric history (**Table 1**).

32% of our population were diagnosed with HIV during their current pregnancy, and most of the cases they believed that it was sexually transmitted (**Table 2**).

**Table 2.** Infectious history and ART management of women living with HIV in postpartum

| Characteristic | Mean±SD [range]<br>Frequency (%)<br>N=75 |
| --- | --- |
| Other chronic diseases |  |
| Any | 6 (8) |
| None | 69 (92) |
| Perceived HIV transmission route |  |
| Sexual relationships | 64 (85) |
| Needles | 3 (4) |
| Blood transfusion | 3 (4) |
| Vertical | 2 (3) |
| Do not know | 3 (4) |
| Years since the HIV diagnosis (years) | 6±5 [0-25] |
| 0 to 2 | 24 (32) |
| 3 to 5 | 18 (24) |
| 6 to 10 | 20 (27) |
| 11 to 15 | 9 (12) |
| 16 to 20 | 3 (4) |
| More than 20 | 1 (1) |
| HIV Symptoms |  |
| Asymptomatic | 73 (97) |
| Symptomatic | 2 (3) |
|  | n=71 |
| Last CD4 count during pregnancy (cells/mm<sup>3</sup>) | 471±253 [36-1,162] |
| More to 500 cells/mm<sup>3</sup> | 31 (41) |
| 200 to 499 cells/mm<sup>3</sup> | 28 (37) |
| Less to 200 cells/mm<sup>3</sup> | 12 (16) |
|  | n=71 |
| Initial viral load (copies/mL) | 17,869±31,217 [41-13'000,095] |
| Undetectable | 41 (58) |
| Less 400 copies/mL | 4 (6) |
| 401 to 100 mil copies/mL | 25 (35) |
| More 100 mil copies/mL | 1 (1) |
| Previous HIV-infected children | n=74 |
| Yes | 7 (9) |
| No | 67 (91) |
| Current partner HIV-status | n=66 |
| Healthy | 31 (47) |
| HIV+ | 18 (27) |
| Do not know | 17 (26) |
| ART initial | n=74 |
| Group a | 17 (23) |
| Group b | 8 (11) |
| Group c | 49 (66) |
| Adherence to ART |  |
| High | 63 (84) |
| Medium | 5 (7) |
| Low | 7 (9) |

Table 2. (continued)
| Characteristic | Frequency (%)<br>n=75 |
| --- | --- |
| Intrapartum prophylactic ART |  |
| Yes | 58 (77) |
| No | 17 (23) |
| Efavirenz during pregnancy |  |
| Yes | 18 (25) |
| No | 53 (75) |
| Perception of perinatal complications | n=74 |
| Yes | 26 (35) |
| No | 48 (65) |

SD: Standard deviation. ART: Antiretroviral therapy. ART Group a: 2 Nucleoside Reverse Transcriptase Inhibitors (NRTIs) + 1 Non- Nucleoside Reverse Transcriptase Inhibitor (NNRTI). ART Group b: 2 NRTIs + 1 Protease Inhibitor (PI). ART Group c: 2 NRTIs + 1 Integrase Inhibitor (INI).

All participants received ART immediately after the diagnosis and during their pregnancies with effective clinical management, as indicated by CD4+ levels and undetectable viral loads, preventing vertical transmission (**Table 2**). The most common ART regimen included two reverse transcriptase inhibitors plus integrase inhibitors (63%) categorized as group C (**Table 2**). According to the national guidelines for the care of WLWV [26], during birth, 76% of participants received ART intrapartum as prophylaxis. Based on questionnaires applied during postpartum period, the adherence to ART perception was high (84%), though one in three women reported perinatal complications.

### Quality of life in the Postpartum-WLWH

In our study population, the WHOQoL-HIV-BREF instrument demonstrated excellent reliability (Cronbach’s index, α= 0.8). The total QoL score was 15±2.0, with the physical health and level of independence domains scoring highest (16±1.2) and the psychological health domain scoring lowest (14±2.0) (**Table 3**).

**Table 3.** WHOQOL-HIV BREF instrument scores of women living with HIV in postpartum

| *Domains*<br>*Subdomains* | *WHOQoL-HIV Bref score*<br>*transformed 4-20*<br>*mean ±SD* |
| --- | --- |
| Overall QoL and General health | 15.5 (±3.0) |
| Quality of Life | 15.5 (±2.6) |
| Health status | 15.6 (±3.4) |
| I. Physical Health | 16 (±1.20) |
| Pain and Discomfort | 14.8 (±4.3) |
| Symptoms of WLWH | 15.6 (±5.4) |
| Energy and fatigue | 14.7 (±3.8) |
| Sleep and rest | 16.1 (±3.4) |
| II. Psychological Health | 14 (±2.0) |
| Positive feelings | 16.2 (±5.4) |
| Cognitive skills | 12.9 (±6.2) |
| Bodily image and appearance | 15.5 (±3.6) |
| Self-esteem | 15.9 (± 3.5) |
| Negative feelings | 12.1 (±4.3) |
| III. Level of Independence | 16 (±0.7) |
| Dependence of medication or treatments | 15.4 (±6.0) |
| Mobility | 16.6 (±3.0) |
| Activities of daily living | 15.3 (±2.8) |
| Work capacity | 15.6 (±3.6) |
| IV. Social Relationships | 15 (±1.20) |
| Social inclusion | 16.5 (±3.7) |
| Personal relationships | 14.6 (±3.4) |
| Sexual activity | 13.4 (±4.6) |
| Social support | 14.6 (±4.3) |
| V. Environment | 14 (±2.4) |
| Physical safety and security | 14.8 (±3.5) |
| Home environment | 14.8 (±3.8) |
| Financial resources | 10.7 (±3.9) |
| New information and skills | 16.5 (±4.3) |
| Recreation and leisure activities | 10.8 (±5.0) |
| Physical environment | 15.4 (±4.0) |
| Health and social care: accessibility and quality | 17.2 (±2.6) |
| Transport | 13.1 (±4.4) |
| VI. Spirituality, Religion and Personal beliefs | 14 (±2.4) |
| Sense of life (SRPB) | 17.0 (±3.5) |
| Forgiveness and blame | 11.8 (±7.1) |
| Concerns about future | 12.6 (±5.3) |
| Death and dying | 12.3 (±6.6) |
| Total quality of life (QoL) | 15 (±2.0) |

Financial resources, recreation and leisure activities, forgiveness and blame, negative emotions, future concerns, death and dying, and cognitive skills were the sub-domains with the lowest scores (**Table 3**). A lack of adequate funds probably made it harder to find time for relaxation and pleasure, which meant that worries and pessimistic thoughts went unaddressed.

Using a cut-off point of 15±2, QoL classifications were defined as poor (<15), medium (15), and high (>15). Despite an overall medium QoL, 49% of WLWH perceived a poor total QoL (**Table 4**). **S1 Table** indicates that the QoL score was not significantly influenced by any of the three antiretroviral therapy groups.

**Table 4.**
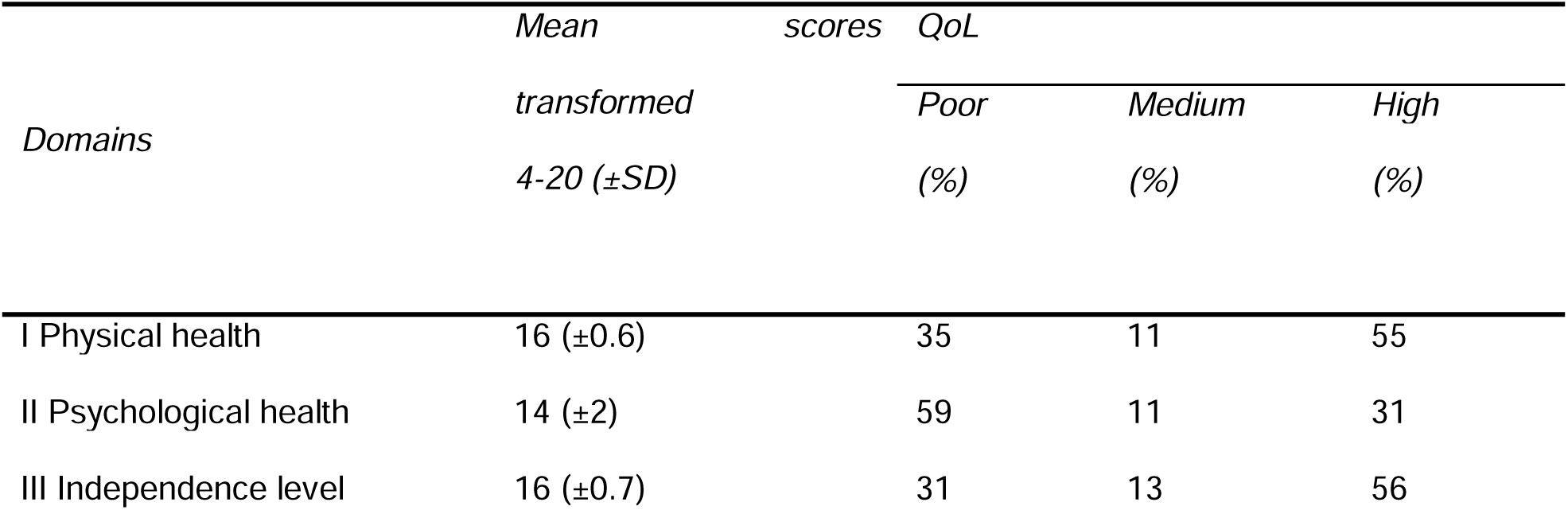
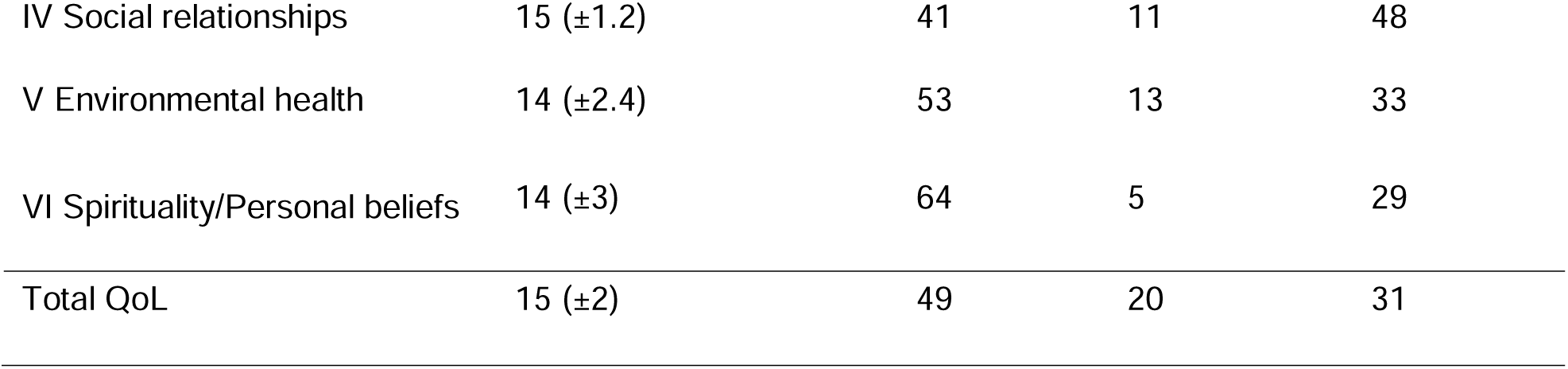
Percentage of women living with HIV in postpartum with poor QOL scores

| *Domains* | *Mean* | *scores* | *QoL* |  |  |
| --- | --- | --- | --- | --- | --- |
|  | *transformed* |  | *Poor* | *Medium* | *High* |
|  | *4-20 (±SD)* |  | *(%)* | *(%)* | *(%)* |
| I Physical health | 16 (±0.6) |  | 35 | 11 | 55 |
| II Psychological health | 14 (±2) |  | 59 | 11 | 31 |
| III Independence level | 16 (±0.7) |  | 31 | 13 | 56 |

|  |  |  |  |  |
| --- | --- | --- | --- | --- |
| IV Social relationships | 15 ($\pm 1.2$) | 41 | 11 | 48 |
| V Environmental health | 14 ($\pm 2.4$) | 53 | 13 | 33 |
| VI Spirituality/Personal beliefs | 14 ($\pm 3$) | 64 | 5 | 29 |
| Total QoL | 15 ($\pm 2$) | 49 | 20 | 31 |

About postpartum duration, most of the participants had less than a year since their last obstetric event. There were not significantly differences in the QoL between postpartum time groups (**S2 Table**). The QoL scores of this group of postpartum-WLWH might have been affected because they were enrolled during the COVID-19 shutdown. The physical health domain, which addresses issues including pain, discomfort, HIV symptoms, rest, and sleep, notably decreased during this time, with the lowest ratings occurring in 2022 (Kruskal-Wallis, *p*= 0.0375) (**S2 Table**).

### CD4 T cells related to QoL in postpartum-WLWH

CD4 count was significantly correlated with four domains. Physical health (r_s_=0.23, *p*=0.04), Psychological health (r_s_=0.027, *p*=0.01), independence (r_s_=0.025, *p*=0.03), and social relationships (r_s_ =0.34, *p*=0.003) as shown in **Table 5**. Other significant correlation included monthly income and psychological health (r_s_=0.26, p=0.04), year of the interview with physical health (r_s_=-0.37, *p*=<0.01) and the application of tattoos or piercings with the personal beliefs (r_s_= -0.32, *p*=0.01) (**Table 5**).

**Table 5.** Association among the QOL domains scores and demographic-clinical variables of women living with HIV in postpartum

|  | *Physical health* | *Psychological health* | *Independence level* | *Social relationships* | *Environment* | *Personal beliefs* |
| --- | --- | --- | --- | --- | --- | --- |
|  | *rho* | *rho* | *rho* | *rho* | *rho* | *rho* |
| Last CD4 count during pregnancy | **0.2392\*** | **0.2789\*** | **0.2534\*** | **0.3427\*\*** | 0.1854 | 0.1903 |
| Age of the beginning of sexual life | 0.1171 | **0.3420\*\*** | 0.1646 | 0.0506 | 0.0406 | **0.3250\*** |
| Year of interview | **-0.370\*\*** | 0.0239 | -0.1919 | 0.0581 | -0.1673 | -0.0567 |
| Other chronic diseases | **-0.3319\*\*** | 0.1075 | -0.1545 | 0.0553 | 0.1313 | -0.1096 |
| Monthly income | 0.0759 | **0.2632\*** | 0.1144 | 0.1998 | 0.1744 | 0.0404 |
| HIV symptoms | -0.0866 | -0.2269 | **-0.2292\*** | -0.1840 | -0.1458 | -0.0940 |
| Number of sexual partners | -0.2217 | -0.2321 | **-0.4206\*\*\*** | -0.1020 | 0.0080 | 0.0014 |
| Gestations | 0.0647 | 0.2210 | **0.2817\*** | 0.1014 | -0.1943 | 0.2254 |
| Current birth Delivery | -0.1003 | -0.0656 | 0.0054 | **-0.2846\*** | -0.1482 | -0.0390 |
| Sexual abuse history | 0.2271 | 0.0453 | 0.0704 | **0.2355\*** | 0.2254 | 0.0588 |
| Tattoo and piercings | -0.239 | -0.2535 | -0.1687 | 0.0818 | 0.06501 | **-0.3210\*** |

*rho*: Spearman correlation test, *p* value is significant when it is <0.05. ART: Antiretroviral therapy. QoL:
Quality of life. WLWH: Women living with HIV.
\**p*<0.05
\*\**p*<0.01
\*\*\**p*<0.001

ART schemes showed no domain correlation though Efavirenz-based regimens, were negatively and significantly associated with Physical health (r_s_= -0.30, *p*=0.01) and independence (r_s_= -0.24, *p*=0.04) (**S3 Table**). Independence correlated negatively with HIV symptoms (r_s_= -0.23, *p*=0.04), while physical health correlated negatively with comorbidities (r_s_= -0.33; p <0.01). Social relationship correlated negatively with the route of birth (r_s_= -0.28, *p*=0.01). Initiation of the sexual life correlated positively with psychological health (r_s_=0.34, *p*<0.01) and personal beliefs (r_s_=0.32, *p*=0.01) Independence was negatively correlated with number of sexual partners (r_s_= -0.42, *p*<0.001) but positively with the number of gestations (r_s_=0.28, *p*=0.01). Social relationships showed a positive correlation with sexual abuse history (r_s_=0.24, *p*=0.04). Additional factors influencing HIV are detailed in **S3Table**.

## Discussion

The UNAIDS 2030 95-95-95 targets do not include the ’fourth 95’ target officially, which envisions that 95% of people living with HIV (PLWH) achieve a QoL comparable to individuals living without HIV. This target emphasises the identification of factors that reduce QoL and the establishment of strategies to address them and support progress toward its achievement [4–8].

Our participants received medical care at a tertiary medical institution in Mexico City, our findings align with previous studies [27–34], showing that HIV/SIDA disproportionally affects the population in disadvantaged socioeconomic strata. Limited education restricts access to well-paid employment. These women live in marginalised housing areas around Mexico City, lacking essential services like transport and security. This condition results in discrimination, gender and social inequality, and economic dependence (**Table 1**).

Most participants received ART (2NRTI + 1 II) during their pregnancies; they perceived high adherence, which thrived until postpartum, as reported in their clinical file. Many also received intrapartum prophylactic ARTs according with the national guidelines for the care of WLWV [26], and underwent Caesarean sections (**Table 2**). Studies indicate a high adherence to ART during pregnancy, which decreases after delivery [30]. Pregnant WLWH prioritise adherence to prevent vertical transmission and ensure their health to care for their babies [31]. Most of the participants had a recent diagnosis, which may help patients remain physically well and focus on their ART, despite psychological distress [11].

In our sample, 47% of the couples were serodiscordant, (**Table 2**) and disclosed their diagnosis only to current partners or close family, no extended relatives. This behaviour coincides with previous studies, where PLWH only tell their partners or immediate family members about their diagnosis. Extended family members, such as parents-in-law, siblings, and grandparents, are not informed due to stigmatization [35,36]. Parental support from partners during pregnancy has been linked to better adherence to ART [37].

The QoL instrument demonstrated excellent internal consistency. In agreement with the findings of the English version in Ethiopia [28,32], as well as the Spanish version [38].

The Mexican postpartum WLWH group had an intermediate mean total QoL score (**Tables 3 and 4**) despite belonging to the lowest socioeconomic strata. Their undetectable viral load, adequate CD4 cell count, and adherence to ART contributed to an optimistic perception and HIV-free births. Nevertheless, the monthly family income influenced the QoL (**Table 5**), reflecting the impact of socioeconomic constraints despite the high Human Development Index (HDI) of Mexico (HDI, 0.781) [39]. Vulnerable groups face limited access to essential services, lowering QoL scores [38]. A study in India [27], found higher QoL scores, despite the poverty of the non-pregnant-WLWH, attributing this to a structured programme for the distribution of ART, close monitoring of adherence, and strong community support, considering that India has a lower HDI (0.644) compared with Mexico. In contrast, in Ethiopia (HDI, 0.492), Iran (HDI, 0.780), Brazil (HDI, 0.760), and even Spain (HDI,0.911) [39], limited education, lack of community support, barriers to ART access due to gender, socioeconomic inequalities, stigma, and racism were related to poor QoL, low adherence, and severe health problems [28,29,34,38]. We suggest that HDI is not associated with the perception of QoL.

The total QoL comprises six domains (**Table 4**) with global differences most evident in mean scores. Our participants reported energy for daily tasks (physical and independence level domains). And satisfaction with their relationships due to partners, friends, or family support (relationship domain). However, the stigma surrounding their condition often led to secrecy, anxiety, and reduced QoL scores in the domains of psychological health and personal beliefs. According to a study in northern Mexico, stigma and a lack of societal acceptance of people living with HIV/AIDS (PLWH) discourage them from disclosing their status to family, friends, and coworkers [40].

Our postpartum-WLWH expressed dissatisfaction with their living conditions due to insufficient services and transportation, in addition to financial constraints that limit access to health care and leisure activities, as reflected in a low environmental domain score (**Table 3**). Moreover, their wages were lower than men’s when they were employed; likewise, the low psychological health scores highlight ongoing stigma and discrimination, with perception of judgment tied to forgiveness and guilt, as reported in Spain [38], Ethiopia [28,32], Iran [33], Brazil [29], and India [27]. Also, we could say that the QoL perceptions varied according to cultural, traditions, and religious factors. In this respect, Haraldstat *et al*. concluded that the concept of QoL may vary across cultures, with unclear cross-cultural relevance. Further studies in Asian and non-Western cultures are needed to explore QOL and its cultural manifestations [18].

Our findings showed that CD4 cell count influenced four of six QOL domains (**Table 5**), the acceptable counts that could reduce their perception of fear, anxiety and may improve well-being for postpartum WLWH and their children, increasing their QOL in many domains. Consistent with previous research [27,38], acceptable levels of CD4 cells from adherence to ART are associated with better perception of QoL.

The global population experienced significant impacts from the SARS-CoV-2 (COVID-19) pandemic, including economic, social, psychological, environmental, and public health challenges. The disease posed life-threatening risks and immense psychological stress worldwide. The relationship between the perception of vulnerability (e.g., unemployment, food, job insecurity, and illness) and low QoL is widely accepted [41]. Teotonio *et al*. reported that females presented worse QoL than males, due to predominantly manage family food purchases, meal preparation, and household decisions. During social distancing, women bore most domestic chores, childcare, education, and professional responsibilities, adversely affecting their QoL [41]. Mexican postpartum WLWH were not the exception, they had reduced scores in the physical health domain in 2022 compared to 2020-2021 scores (**Table 5**). In accordance with other studies [41–47] also the increased stress, isolation, gender inequities, and reduced physical activity during lockdown, exacerbated vulnerability and wellness perceptions among WLWH.

No single instrument can fully evaluate all QoL factors. WHOQOL-HIV-BREF, although robust, does not fully assess barriers, such as mental illness and addictions or specific aspects related to ART (e.g., adherence, duration and type). More research should use specialised tools to assess adherence to ART and mental health. It is important to stablish that this instrument assesses the QoL and include two questions about self-perceived QoL and health status, which score we called overall QoL and general health (**Table 3**).

This study had several limitations. First, obtaining a large sample of women living with HIV (WLWH) was challenging due to the specificity of their characteristics and vulnerability. Second, the cross-sectional design prevented the establishment of causality between variable associations. Third, ART adherence was obtained through clinical interviews rather than specialized instruments. Fourth, quality of life (QoL) behavior may have been influenced by the lockdown and post-pandemic period of the SARS-CoV-2 (COVID-19) pandemic.

Despite its limitations, this study provides valuable information on highly vulnerable populations such as postpartum WLWH and their QOL scores. The sample size is representative of the Mexican population because the study was conducted at a tertiary medical institution. It emphasises identifying and addressing the factors affecting the perception of QoL to propose strategies for medical attention and therapeutic success, overall well-being in PLWH.

Advocate for including QoL evaluation in WHO strategies, the fourth 95 target, and government and society efforts to reduce stigma, foster acceptance, and improve environments for WLWH, future generations, including exposed newborns.

## Conclusions

HIV/AIDS negatively affects the QoL of postpartum WLWH and potentially their babies. Despite poverty, stigma, and discrimination, they remain motivated, especially when their child is born HIV-free. They perceived their QoL as moderate, hindered by low-income. Clinical factors, particularly optimal CD4 cell count through ART and high adherence, which are closely linked to improved QoL. These findings emphasise the importance of formally including QoL assessment as the fourth target of the ’95’ in WHO strategies for PLWH.

## Acknowledgments

The authors are also deeply grateful to all the women who participated in this work, and we hope that our efforts will improve their quality of life in all respects. The authors thank to INPer (approval number 212250-3120771), as well as their colleagues, Estela Y. Godínez-Martínez, Virginia E. Santillán-Palomo, María del Pilar Meza-Rodríguez, Carmen Flores-Cisneros, Cecilia Mota-González, Rosa Georgina Rodríguez-Delgado, and Edgar Bonilla-Reyes, for their excellent technical assistance, interest in this work, and helpful discussions.

## Supporting information

**S1 File.** WHOQoL-HIV BREF instrument (Spanish version)

**S1 Table**. Comparison of mean scores of domains for QOL in postpartum-WLWH using three different combinations of ART

**S2 Table.** QOL scores by domains in different postpartum times and years of interviews of postpartum-WLWH

**S3 Table.** Association among the QOL domain scores and sociodemographic-clinical variables in postpartum-WLWH

## Authors’ Contributions

**Conceptualization** by Hernández-Pineda J. **Data curation** was performed by Domínguez-Castro M, Ramírez-Ramírez A, Avila-García M, Hernández-Pineda J **Formal analysis** was performed by Ramírez-Ramírez A, Domínguez-Castro M, Avila-García M, López-Martínez M, Hernández-Pineda J, Mancilla-Herrera I, and Figueroa-Damián R. **Investigation** was performed by Domínguez-Castro M, Ramírez-Ramírez A, Plazola-Camacho NG, Figueroa-Damián R, Hernández-Pineda J, Romo-Yáñez J, and García-Romero CS **Methodology** was performed by Ramírez-Ramírez A, Domínguez-Castro M, Hernández-Pineda J, Plazola-Camacho NG, López-Martínez M, Mancilla-Herrera. **Project administration** was performed by Soriano-Becerril, DM, and Figueroa-Damián, R. Medical attention was provided by Plazola-Camacho, NG. **Supervision** of the project was performed by Ramírez-Ramírez A, Domínguez-Castro M, Plazola-Camacho N, Romo-Yáñez J, Katende-Kyenda NL, and Hernández-Pineda J. **Validation** was performed by Domínguez-Castro M, Hernández-Pineda J, Avila-García M, Katende-Kyenda NL, Soriano-Becerril DM, García-Romero CS, López-Martínez M and Figueroa-Damián R. **Visualization** was performed by Ramírez-Ramírez A, Domínguez-Castro M, Hernández-Pineda J, Plazola-Camacho NG, López-Martínez M, Mancilla-Herrera, and Romo-Yáñez J. **Writing**-**Original draft preparation** by Ramírez-Ramírez A, Domínguez-Castro M, Avila-García M, and Hernández-Pineda J**. Writing-Review and editing** of the manuscript were performed by all authors.

## Ethical statement

This study was conducted according with the ethical standards set by the Institutional and National Research Committees, and the 1964 Declaration of Helsinki and its later amendments. All women provided informed consent prior to their involvement in the research. The study protocol was reviewed and approved by the INPer Ethics Committee; and all data collected were treated with confidentiality and used solely for this research.

## Data Availability Statement

The data that support the findings of this study are available from the corresponding author upon reasonable request.

