## Supplementary material for "Quality of life associated with clinical and sociodemographic determinants in the postpartum period of Mexican women living with HIV. A cross-sectional study": S1. Table

| **S1 Table.** Comparison of mean scores of domains for QOL in postpartum-WLWH using three different combinations of ART | | | | | |
| --- | --- | --- | --- | --- | --- |
| *Domains* | *ART* | | | | |
|  | *Group A*  *Mean ±SD*  *n= 17* | | *Group B*  *Mean ±SD*  *n= 8* | *Group C*  *Mean ±SD*  *n= 49* | *P value^a^* |
| I Physical health | | 16 (±1.2) | 17 (±1.7) | 15 (±0.7) | 0.2240 |
| II Psychological health | | 16 (±2.0) | 14 (±4.0) | 15 (±2.0) | 0.7816 |
| III Independence level | | 16 (±0.7) | 17 (±0.7) | 15 (±0.2) | 0.1419 |
| IV Social relationships | | 15 (±0.8) | 15 (±1.0) | 14 (±0.4) | 0.6152 |
| V Environment | | 15 (±1.9) | 14 (±2.2) | 14 (±2.4) | 0.9162 |
| VI Spirituality/Personal beliefs | | 15 (±2.8) | 11 (±2.1) | 13 (±2.3) | 0.1265 |
| Total QoL | 16 (±1.6) | | 15 (±2) | 14 (±1.3) | 0.1278 |

ART: Antiretroviral therapy. QoL: Quality of life. SD: Standard deviation. Group a: 2 nucleoside reverse transcriptase inhibitors (NRTIs) + 1 nonnucleoside reverse transcriptase inhibitor (NNRTI). Group b: 2 NRTIs + 1 protease inhibitors (PI). Group c: 2 NRTIs + 1 integrase inhibitors (INI).

a: Kruskal‒Wallis test, *p* value is significant when is <0.05.
