## Supplementary material for "Quality of life associated with clinical and sociodemographic determinants in the postpartum period of Mexican women living with HIV. A cross-sectional study": S2. Table

**S2 Table.** QoL scores by domains in different postpartum times and years of interviews of postpartum-WLWH

| *Domains* | *Pospartum times* | | | | *Years of interviews* | | | |
| --- | --- | --- | --- | --- | --- | --- | --- | --- |
|  | *Less 1 year*  *n=55* | | *Over 1 year*  *n=20* | *P-value^a^* | *2020*  *n= 20* | *2021*  *n= 49* | *2022*  *n= 6* | *P-value^b^* |
| I Physical health | | 15±0.9 | 16±1.6 | 0.2000 | 16±0.5 | 16±1.1 | 10±4.4 | **0.0375** |
| II Psychological health | | 15±2.0 | 15±2.0 | 0.9142 | 15±1.9 | 15±1.5 | 16±1.5 | 0.8442 |
| III Independence level | | 16±0.9 | 17±0.1 | 0.2425 | 16±0.6 | 16±0.6 | 13±3.4 | 0.3819 |
| IV Social relationships | | 15±1.3 | 15±0.9 | 0.5590 | 14±1.8 | 15±1.2 | 15±2.1 | 0.7574 |
| V Environment | | 14±2.7 | 14±3.0 | 0.8743 | 17±1.1 | 16±1.1 | 16±1.4 | 0.6461 |
| VI Spirituality/Personal beliefs | | 14±2.3 | 13±3.1 | 0.7715 | 12±3.8 | 14±1.8 | 12±4.5 | 0.3792 |
| Total QoL | 15±1.7 | | 15±1.8 | 0.8669 | 15±1.5 | 15±0.8 | 14±2.7 | 0.4478 |

ART: Antiretroviral therapy. SD: Standard deviation. WLWH: Women living with HIV.

^a^ U Mann-Whitney’s test, *p*-value <0.05.

^b^ Kruskal-Wallis test, *p*-value <0.05.
