## Supplementary material for "Quality of life associated with clinical and sociodemographic determinants in the postpartum period of Mexican women living with HIV. A cross-sectional study": S3. Table

**S3 Table.** Association among the QOL domain scores and sociodemographic-clinical variables in postpartum-WLWH

|  | *Physical health* | *Psychological health* | *Independence level* | *Social relationships* | *Environment* | *Personal beliefs* |
| --- | --- | --- | --- | --- | --- | --- |
|  | *rho* | *rho* | *rho* | *rho* | *rho* | *rho* |
| *Demographic variables* | | | |  |  |  |
| Age | 0.1123 | 0.1713 | 0.1666 | 0.1958 | -0.0305 | 0.1247 |
| Gestational age at medical admission | 0.1009 | -0.0119 | 0.0813 | -0.1645 | -0.0413 | 0.0638 |
| Educational status | -0.1043 | -0.1158 | -0.2236 | 0.1187 | 0.0630 | -0.0464 |
| Marital status | 0.1663 | 0.0654 | 0.1234 | 0.0327 | -0.0609 | -0.1602 |
| Employment status | -0.1210 | -0.0198 | -0.1245 | -0.1604 | -0.1130 | 0.2002 |
| Place of residence | -0.0478 | 0.1271 | 0.1238 | 0.0246 | 0.1082 | 0.0011 |
| Place where they come from | -0.0532 | 0.0590 | -0.0222 | -0.1922 | -0.2185 | 0.0462 |
| Monthly income | 0.0759 | **0.2632*** | 0.1144 | 0.1998 | 0.1744 | 0.0404 |
| Year of interview | **-0.370**** | 0.0239 | -0.1919 | 0.0581 | -0.1673 | -0.0567 |
| *Non-pathologic history variables* | | |  |  |  |  |
| Smoking habit | -0.088 | -0.0904 | -0.1452 | -0.1239 | 0.0212 | -0.0912 |
| Alcoholism and Drugs Addiction | -0.158 | -0.1357 | -0.1523 | -0.1178 | -0.0647 | -0.2369 |
| Tattoo and piercings | -0.239 | -0.2535 | -0.1687 | 0.0818 | 0.06501 | **-0.3210*** |
| Partner’s addictions | 0.2217 | -0.2215 | 0.1512 | 0.05512 | 0.0277 | -0.1897 |

*rho*: Spearman correlation test, *p* value is significant when it is <0.05. ART: Antiretroviral therapy. QoL: Quality of life. WLWH: Women living with HIV.

**p*<0.05

***p*<0.01

****p*<0.001

**TABLE S3** (*CONTINUED*)

|  | | Physical health | Psychological health | Independence level | Social relationships | Environment | Personal beliefs |
| --- | --- | --- | --- | --- | --- | --- | --- |
|  |  | *rho* | *rho* | *rho* | *rho* | *rho* | *rho* |
| *Infectious history and ART management* | | | | |  |  |  |
| Other chronic diseases | **-0.3319**** | | 0.1075 | -0.1545 | 0.0553 | 0.1313 | -0.1096 |
| Years living with HIV | 0.0937 | | 0.1464 | 0.1665 | 0.05381 | 0.0800 | -0.0443 |
| HIV symptoms | -0.0866 | | -0.2269 | **-0.2292*** | -0.1840 | -0.1458 | -0.0940 |
| Last CD4 count during pregnancy | **0.2392*** | | **0.2789*** | **0.2534*** | **0.3427**** | 0.1854 | 0.1903 |
| Initial viral load | 0.0221 | | -0.1424 | -0.0501 | -0.2185 | -0.1483 | -0.1953 |
| Previous HIV-infected children | -0.0206 | | -0.1957 | -0.1220 | -0.1260 | -0.0119 | -0.1009 |
| Current partner HIV-status | -0.0909 | | -0.2746 | -0.1000 | -0.2707 | -0.0285 | -0.0511 |
| Schem of ART | -0.1406 | | -0.0347 | 0.8480 | 0.5414 | 0.5935 | 0.1280 |
| Adherence to ART | -0.0157 | | 0.0971 | 0.0506 | -0.0380 | -0.0036 | 0.1347 |
| Intrapartum prophylactic ART | -0.0577 | | 0.1206 | 0.0496 | -0.1416 | -0.0605 | -0.067 |
| ART schem containing efavirenz | **-0.3039*** | | -0.2202 | **-0.2444*** | -0.1218 | -0.0681 | -0.1934 |
| Perception of pregnancy complications | 0.1108 | | 0.0286 | 0.1061 | 0.0812 | 0.2015 | 0.0505 |

*rho*: Spearman correlation test, *p* value is significant when it is <0.05. ART: Antiretroviral therapy.

**p*<0.05

***p*<0.01

****p*<0.001

**TABLE S3** *(CONTINUED)*

|  | Physical health | Psychological health | Independence level | Social relationships | Environment | Personal beliefs |
| --- | --- | --- | --- | --- | --- | --- |
|  | *rho* | *rho* | *rho* | *rho* | *rho* | *rho* |
| *Obstetric history and sexual behaviour* | | | |  |  |  |
| Age of the beginning of sexual life | 0.1171 | **0.3420**** | 0.1646 | 0.0506 | 0.0406 | **0.3250*** |
| Number of sexual partners | -0.2217 | -0.2321 | **-0.4206***** | -0.1020 | 0.0080 | 0.0014 |
| Gestations | 0.0647 | 0.2210 | **0.2817*** | 0.1014 | -0.1943 | 0.2254 |
| Births | 0.2090 | -0.0526 | 0.0867 | -0.0343 | -0.0377 | 0.0706 |
| Caesarean sections | -0.0773 | 0.0349 | 0.0782 | -0.0004 | 0.0086 | -0.0117 |
| Abortions | -0.0514 | -0.0335 | 0.0290 | 0.0570 | -0.1336 | 0.1062 |
| Time of postpartum | 0.1964 | -0.0146 | 0.2169 | -0.0080 | -0.0142 | -0.0598 |
| Current birth Delivery | -0.1003 | -0.0656 | 0.0054 | **-0.2846*** | -0.1482 | -0.0390 |
| Sexual abuse history | 0.2271 | 0.0453 | 0.0704 | **0.2355*** | 0.2254 | 0.0588 |

*r*: Spearman correlation test, *p* value is significant when it is <0.05.

**p*<0.05

***p*<0.01

****p*<0.001
